# Posttraumatic stress and growth in mothers of children with genetic diseases in early childhood: a protocol of a longitudinal study

**DOI:** 10.1101/2025.04.03.25325222

**Authors:** Kołecka Aleksandra, Borchet Judyta, Bieleninik Łucja

## Abstract

**Introduction:** Diagnosing a child’s disease is a traumatic event that impacts parents’ psychological well-being, mental health, and is associated with burden. This protocol of a longitudinal study aims to observe posttraumatic stress levels in mothers of children with a genetic disease. It will also explore posttraumatic growth in these mothers and identify key predictors of such growth.

**Materials and Methods:** The project is a sequential study that integrates both cross-sectional and longitudinal designs. The study will approach mothers of children aged 0 to 3 years who have been diagnosed with a genetic disease by a specialist before their first birthday. Two measurements will be taken with a 6-month interval between the first and second assessments. The study will use a questionnaire-based approach. The research assessment will be conducted using: the Impact of Event Scale-Revised (IES-R), the Posttraumatic Growth Inventory (PTGI), the Zimbardo Time Perspective Inventory (ZTPI), the Parental Burnout Assessment (PBA), the Kansas Inventory of Parental Perceptions (KIPP), and the Family Resilience Assessment Scale (FRAS).

**Discussion:** This study may provide valuable insights into how mothers cope with a child’s genetic disease, both from a time perspective and a parental perspective. It could inform the development of targeted support strategies to help mothers manage the challenges of raising a child with a genetic disease, particularly in areas such as crisis intervention or therapy. We hypothesize that improving the mother’s time perspective could improve the mother’s well-being and thus enhance the overall functioning of the family system and support the child’s development.

## Introduction

### Background

Time perspective refers to how a person perceives and experiences time in their life. The perspective influences how individuals organize their experiences, make decisions, and interpret the world around them. Research by Zimbardo and Boyd (2008) suggests that time perspective is relatively stable, but stressful, uncontrollable, and unpredictable situations can alter a person’s time perspective, often narrowing their focus and making it difficult to maintain a broader view of time (1–4). In some cases, it may even cause individuals to concentrate exclusively on one aspect of their time perspective (past, present, or future). Disease is one such event that can significantly change the way time is perceived (2,5–7).

Many studies have shown that patients with various somatic and mental diseases experience changes in their time perspective over time. The experience of disease significantly impacts the time perspective of patients with conditions such as cancer (3, 8-9, 10-12), multiple sclerosis (13), Alzheimer’s disease (14), renal failure (7), inflammatory bowel disease (15), depression (16), schizophrenia (17), and other mental health disorders (18). Patients often become more focused on a negative past and fatalistic present compared to individuals without health conditions (3,4). Additionally, they frequently experience a narrowing of their future time perspective (14).

Experiencing diseases can alter the time perspective not only of the patient but also of their caregivers and family members. Research has shown that caregivers and families of individuals with chronic diseases often notice significant changes in their perception of time. Life becomes more unpredictable, and daily routines are disrupted. Caregivers frequently feel uncertain and anxious about the future, which can increase perceived stress (19). Family members may experience time pressure and the constant need for support and care, leading to overload and fatigue (20). The stress experienced by parents is especially heightened when a child is diagnosed with a chronic disease (21–29).

### Study Rationale

The rationale for this study is to explore the experience of posttraumatic stress and posttraumatic growth in mothers of children with genetic diseases. Although there is a wealth of research on posttraumatic stress and posttraumatic growth in individuals with chronic disease (1–9), limited attention has been given to how these experiences manifest in parents, particularly mothers of children with genetic diseases. This research will fill a critical gap in understanding how a child’s genetic disease impacts family life and provide evidence-based recommendations for supporting mothers and families coping with this difficult situation.

### Objectives and hypothesis

This study will answer the research question: does the mother’s time perspective change significantly during the experience of a child’s genetic disease? It will help us to understand the mechanism behind these changes and identify the factors that influence them. The variables analyzed will be: the intensity of posttraumatic stress, the level of posttraumatic growth, the mother’s time perspective, parental burnout, the perception of the mother’s experiences related to parenthood, and family resilience. The study seeks to provide valuable insights into how mothers navigate the experience of having a child with a genetic disease, both from a time perspective and a parenting perspective. We hope that this will lead to a deeper understanding of the mothers’ situation, their behaviors, and their attitudes toward time.

The main objective of this study is to verify the time perspective adopted by mothers in the course of experiencing a child’s genetic disease.

Specific objectives are as follows:

1. to explore the phenomenon of posttraumatic stress in the mother of a child with a genetic disease in the context of her time perspective
2. to explore the phenomenon of posttraumatic stress in the mother of a child with a genetic disease from a parental perspective
3. to explore the phenomenon of posttraumatic growth in the mother of a child with a genetic disease in the context of her time perspective
4. to explore the phenomenon of posttraumatic growth in the mother of a child with a genetic disease from a parental perspective
5. to indicate predictors of posttraumatic stress in the mother of a child with a genetic disease
6. to indicate predictors of posttraumatic growth in the mother of a child with a genetic disease

We hypothesize in this study that:

1. The mother’s time perspective changes during the experience of the child’s genetic disease (10– 15).
2. Time perspective is associated with the intensity of posttraumatic stress in the mother of a child with a genetic disease: a negative past time perspective increases the intensity of posttraumatic stress in the mother of a child with a genetic disease; a positive past time perspective decreases the intensity of posttraumatic stress in the mother of a child with a genetic disease; a present hedonistic time perspective decreases the intensity of posttraumatic stress in the mother of a child with a genetic disease; a present fatalistic time perspective increases the intensity of posttraumatic stress in the mother of a child with a genetic disease; a positive future time perspective decreases the intensity of posttraumatic stress in the mother of a child with a genetic disease (12,13,16–19). Traumatic events affect the time perspective, make it difficult to act in the full time horizon, and often limit the perception of time to an increased concentration on one of the time perspectives (12,13,16,19). The disease shapes specific living conditions that can generate changes in the perspective of time perception (10,12,14,15). So far, a significant impact of the experience of illness on the time perspective has been demonstrated in patients with: cancer (13,20–24), multiple sclerosis (25), Alzheimer’s disease (17), renal failure (15), inflammatory bowel disease (18), depression (26), schizophrenia (27), as well as other mental illnesses (28). Typically, people experiencing illness differed from healthy individuals in a greater focus on the negative past and fatalistic present (13,19). A narrowing of the future time perspective was often observed (17).
3. Parental burnout is positively associated with the intensity of posttraumatic stress in the mother of a child with a genetic disease (29–34). There is no scientific evidence for a direct relationship between parental burnout and PTSD in parents of children with chronic illness. However, it is known that parents of children with chronic diseases are at increased risk of parental burnout (29,30,32–34). The intensity of parental burnout has been observed, among others, in parents of children with ASD (35–37), in parents of adults with autism spectrum disorder (36), in parents of children with special needs (38), in parents of children with neurodevelopmental disorders (39), in parents of children with type I diabetes (32), in parents of children with cancer (40), in mothers of children with ADHD (41), in mothers of children with a rare genetic disease (31) and in mothers of children with complex medical problems (42). Parental burnout is a response to long-term parenting stress (37), is associated with physical and emotional exhaustion, and may pose a potential risk for the development of posttraumatic stress disorder.
4. Family resilience is negatively associated with the intensity of posttraumatic stress in the mother of a child with a genetic disease (43–49). Scientific research emphasize the importance of family resilience in the process of effectively coping with stress experienced by parents, including in the case of families with children with mental, emotional and behavioral disorders (44), childhood asthma (43), type 1 diabetes (47), childhood cancer (49), and childhood coronary artery disease (45,46). Family resilience may be a protective factor against the dynamics of posttraumatic stress (48). At the same time, experiencing a difficult situation, such as a child’s illness, poses a threat to family resources and may, in turn, cause a decrease in family resilience. Studies show that most families with children affected by chronic diseases may exhibit low levels of family resilience (50,51).
5. Time perspective is associated with the level of posttraumatic growth in the mother of a child with a genetic disease: a negative past time perspective decreases the level of posttraumatic growth in the mother of a child with a genetic disease; a positive past time perspective increases the level of posttraumatic growth in the mother of a child with a genetic disease; a present hedonistic time perspective increases the level of posttraumatic growth in the mother of a child with a genetic disease; a present fatalistic time perspective decreases the level of posttraumatic growth in the mother of a child with a genetic disease; a positive future time perspective increases the level of posttraumatic growth in the mother of a child with a genetic disease (18,52–57). There is no scientific evidence of a direct relationship between the parent’s time perspective and the level of posttraumatic growth in the parent of a child with chronic disease. However, studies have shown that the future time perspective is one of the predictors of the occurrence of posttraumatic growth in young adults (52). A genetic disease in a child requires the reorganization of everyday life of the entire family system, directing the parent’s activities to provide optimal support in the development of the child with a genetic disease, which is associated with effort spread over a long time perspective. In view of the above, it is assumed that a positively valued future, concentration on long-term goals, and current support activities may turn out to be a factor contributing to the occurrence of posttraumatic growth in the mother of a child with a genetic disease.
6. Family resilience is positively associated with the level of posttraumatic growth in the mother of a child with a genetic disease (43–49,58–60). Resilience allows the family to adapt to various difficulties or traumatic experiences (60). Many studies emphasize the importance of family resilience in the process of adaptation to a child’s illness, including in the case of families with children with mental, emotional and behavioral disorders (44), childhood asthma (43), type 1 diabetes (47), childhood cancer (49), and childhood coronary artery disease (45,46). Family resilience has been shown to support positive ways of coping with a child’s chronic illness and to develop stronger relationships, better family cohesion, and a positive family belief system (58). High family resilience is an important factor for posttraumatic growth (61,62). Family-related resources increase the chances for an upward trajectory of posttraumatic growth (48,59).

## Materials and Methods

A study protocol has been developed in accordance with the SPIRIT guidelines (see annex). The project received ethics approval from The Research Ethics Board at the University of Gdansk (approval number: 50/2024/WNS, approval date: 23 June 2024).

### Study design

The project is a sequential study that integrates both cross-sectional and longitudinal designs. Two measurements are planned, with a 6-month interval between the first and second assessment. The first participant is expected to be enrolled in the study in September 2024, and the last participant in April 2025. Each undergoes a follow-up assessment 6 months after the initial measurement. The second measurement is scheduled to take place between March 2025 and October 2025 (Fig.1).

**Fig. 1.**
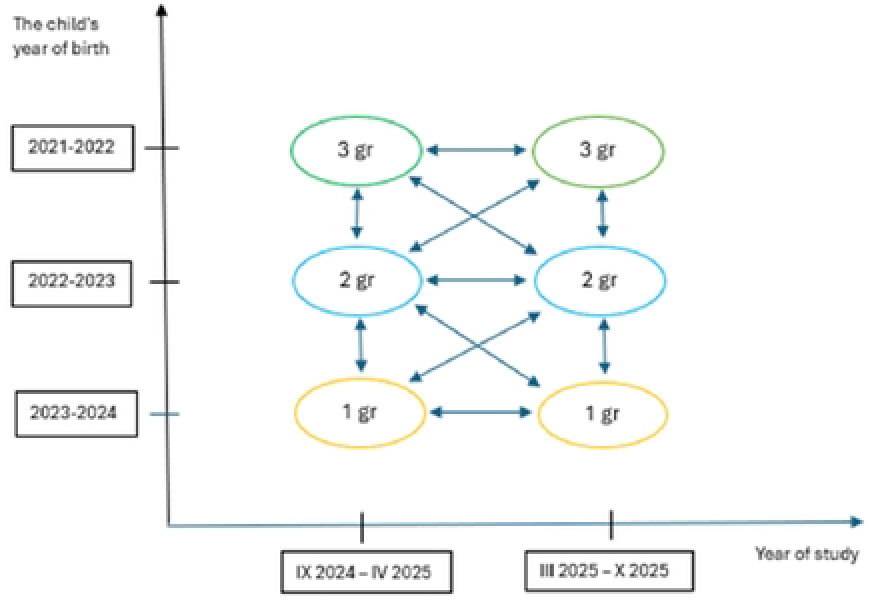
Research design diagram.

### Study procedures

Referrals will be made by an investigator, who will provide both oral and written explanations of the proposed research project. Participants will be informed that enrollment is voluntary and that they may withdraw from the study at any time. Recruitment will be based on the eligibility criteria established for the study group. Each participant will receive detailed information about the study’s objectives and procedures. After obtaining informed consent, an interview will be conducted to collect socio- demographic data. Following this, the mother completes the initial set of questionnaires (baseline assessment).

After the questionnaires are completed, a follow-up meeting will be scheduled approximately six months later. During the second meeting, the mother will complete the same set of questionnaires (2nd assessment) (Fig.2). Each study participant will receive feedback on the study results. Furthermore, each mother will be presented with an opportunity for a psychological consultation with a psychologist experienced in working with people with disabilities and their families.

**Fig. 2.**
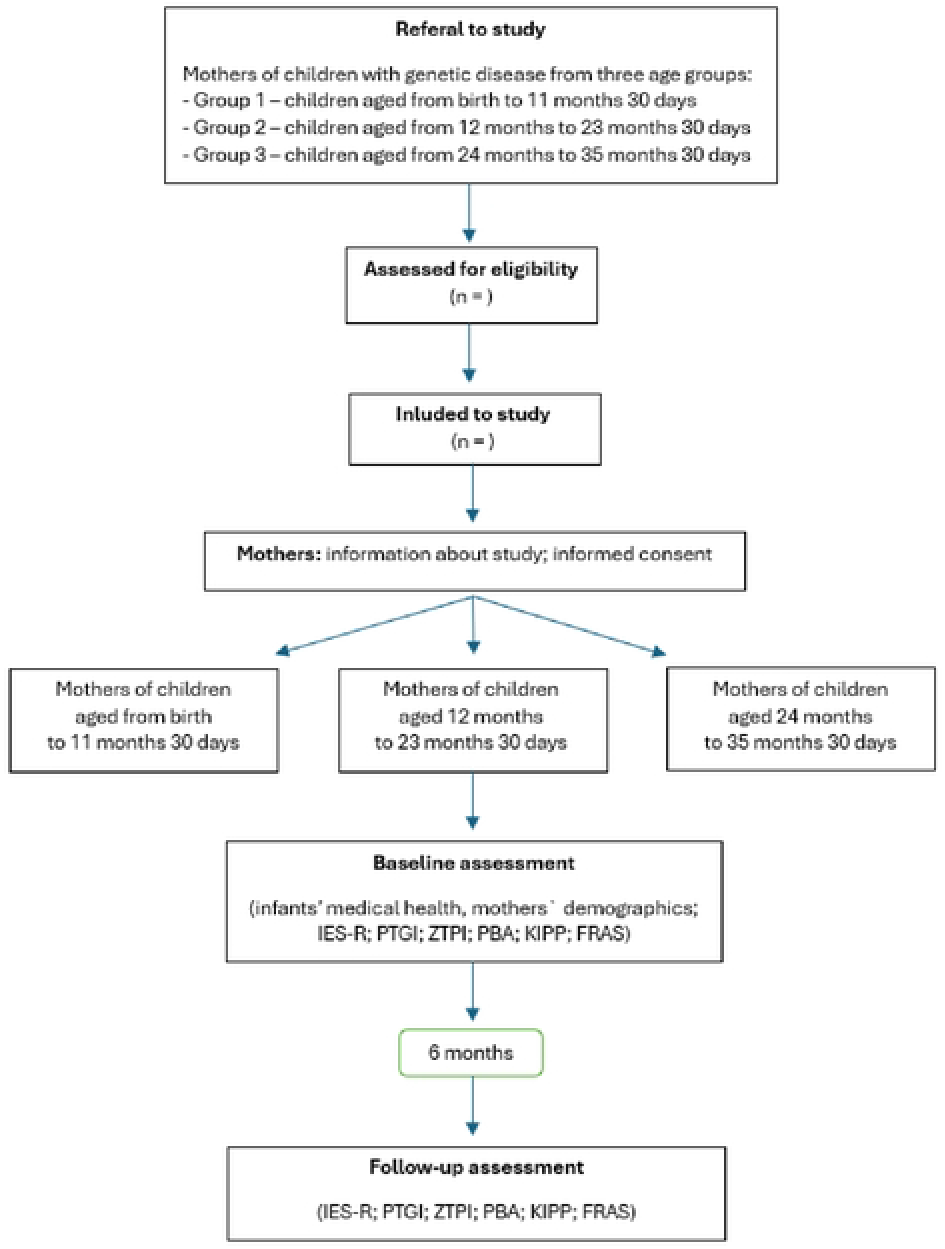
Stages ofthe research. IES-R: Impact of Event Scale Revised (Juczyński & Ogińska-Bulik, 2009) PTGI: Posttraumatic Development Inventory (Ogińska-Bulik & Juczyński, 2010) ZTPI: Zimbardo Time Perspective Questionnaire (Prrepiórka, 2011) PBA: Parental Burnout Questionnaire (Szczygiel et al., 2020) KIPP: Kansas Inventory of Parental Perceptions (Pisula & Noiniska, 2020) FRAS: Family Resilience Assessment Scale (Nadrowska et al., 2021)

The participants will be grouped according to the age of their children into the following categories:

- Group 1 – children aged from birth to 11 months 30 days,
- Group 2 – children aged from 12 months to 23 months 30 days,
- Group 3 – children aged from 24 months to 35 months 30 days.

### Eligibility criteria

#### Inclusion criteria

1. The study will include **mothers**. The rationale for targeting only mothers, rather than fathers, in this study is based on the significant role mothers often play in early child caregiving, especially in supporting children diagnosed with genetic diseases, as mothers usually are their primary caregivers (63–67).
2. **The study will include mothers of children diagnosed with a genetic disease before the age of 1**. The rationale for including only mothers of children diagnosed before the age of 1 year is to allow for meaningful comparisons between the study groups. The criterion of early diagnosis of the child will ensure that each mother will experience the diagnosis of the child’s genetic disease at a similar time in the child’s life. This will make the groups more homogeneous in terms of the timing of the critical event. This limitation will take into account the initial stages of adaptation to the genetic diagnosis, ensuring that the results reflect the unique challenges and stressors experienced by mothers in the time since their child’s diagnosis.
3. The child should be diagnosed by a specialist physician with one of the following genetic diseases: cystic fibrosis (CF), spinal muscular atrophy (SMA), or other genetic disease. The motivation for focusing on these diagnoses stems from consultations with medical personnel. Among all congenital diseases detected in Poland through prenatal testing of newborns, only CF and SMA are genetically determined. In contrast, other conditions may be temporary diagnoses (such as congenital hypothyroidism, hearing defects). Additionally, some congenital metabolic disorders, like phenylketonuria and biotinidase deficiency, can often be managed with appropriate treatment strategies and elimination diets, which may mitigate or eliminate symptoms of the disease.
4. The child must be **under the age of 3**. The rationale for including children up to 3 years of age is based on the critical developmental period that occurs during the first three years of life. This stage is characterized by rapid physical, cognitive, and emotional growth, during which children often experience the initial impact of a genetic disease. The early years are also a time when mothers experience the novelty of this difficult situation, are highly involved in the care of a child with a genetic disorder, and are coping with the challenges of their health. Furthermore, taking into account the periodization of child development and limiting the age of the child to early childhood will ensure better generalization of the results (68,69).

#### Exclusion criteria

1. Having another child with a genetic or chronic disease born earlier. We have decided to exclude parents who have more than one child with a genetic disease or a chronic disease due to previous experiences. Mothers in these situations may have already developed coping strategies for dealing with such traumatic events. This could result in different adaptive patterns, which may not be representative of the initial response to a child’s first genetic diagnosis. As a result, these mothers might exhibit a different intensity of posttraumatic stress or a changed level of posttraumatic growth. Research suggests that repeated exposure to the same type of traumatic event can lead to increased psychological stability and changes in mental resilience, as well as variations in the intensity of post-traumatic stress (30). Additionally, a mother’s perception of time may shift based on the experience gained from previous crisis situations (6).
2. Diagnosis of lethal developmental defects in a child. Mothers of children diagnosed with lethal defects were excluded from the study due to the unique nature of the situation, where the child’s disease ultimately leads to death. The death of a child poses a significant threat to the parenting role and requires specialized support (70). Parenting during a child’s terminal disease involves interventions tailored to this specific experience, which focus on helping parents cope with the impending loss. These interventions include psychological support aimed at acknowledging the parenthood experience, creating positive memories with the child, preparing for the child’s death, and helping parents adjust to a new family dynamic after the loss (71). The diagnosis of lethal defects often requires palliative care interventions (33).

### Measures

1. **The Intensity of the Posttraumatic Stress Symptoms** in mothers will be measured using the Revised Impact of Events Scale (IES-R, Weiss & Marmar, 1997; in the Polish adaptation: Juczyński & Ogińska-Bulik, 2009). The scale consists of 22 statements that assess stress symptoms over the past seven days in relation to the experienced traumatic event. It evaluates three dimensions of posttraumatic stress: Intrusion, Arousal, and Avoidance. Responses are rated on a five-point Likert scale (from “0” - “not at all” to “4” - “definitely yes”). Scores are calculated for the overall scale and for individual dimensions. The results reflect the intensity, rather than the frequency of symptoms experienced by the participant. The cut-off value is set at 1.5 points, which applies to both the general PTSD score and its individual dimensions. Scores exceeding this cut-off value indicate at least moderate posttraumatic stress. A higher score corresponds to greater severity of PTSD symptoms. The tool has achieved satisfactory psychometric properties with internal consistency (Cronbach’s alpha) of .92 for the overall scale, and .89, .85, and .78 for Intrusion, Arousal, and Avoidance —, respectively (34).
2. **The level of the post-traumatic growth** will be measured using the Posttraumatic Growth Inventory (PTGI, Tedeschi & Calhoun, 1996; in the Polish adaptation: Ogińska-Bulik & Juczyński, 2010). The inventory consists of 21 statements that describe positive changes resulting from a negative and traumatic life event. Participants rate each statement on a six-point scale, from “0” (indicating no change as a result of the crisis) to “5” (indicating a very large extent of change). The inventory assesses four factors of posttraumatic growth: Changes in self-perception, Changes in relationships with others, Greater appreciation of life, Spiritual changes. An overall score and individual factor scores are calculated. The overall score is the sum of all the above-mentioned factors with higher scores indicating greater positive change and, therefore, a higher level of posttraumatic growth. The tool demonstrates satisfactory psychometric properties with a Cronbach’s alpha coefficient of .93 for the overall scale and individual factors ranging from .63 to .87) (35).
3. **The time perspective** will be measured using the Zimbardo Time Perspective Inventory (ZTPI, Zimbardo & Boyd, 1999; in the Polish adaptation: Przepiórka, 2011). The inventory consists of 56 statements and assesses the attitude towards time across five dimensions: Present hedonism, Present fatalism, Positive past, Negative past, and Future. Participants rate their agreement with each statement on a five-point scale, where “1” means “I completely disagree” and “5” means “I completely agree”. Scores are calculated for each dimension, reflecting the individual’s subjective attitude toward each time perspective. All scales demonstrate satisfactory internal reliability with Cronbach’s alpha coefficient for individual dimensions as follows: Negative past (α = .84); Positive past (α = .80); Present hedonism (α = .82); Present fatalism (α = .70); Future (α = .82) (36–37).
4. **The parental burnout** will be measured using the Parental Burnout Questionnaire (PBA, Roskam et al., 2017; in the Polish adaptation: Szczygieł et al., 2020), which consists of 23 questions assessing four dimensions of parental burnout: Exhaustion with the parental role, Contrast with the previous image of oneself as a parent, Loss of pleasure in being a parent/Saturation with the parental role, and Emotional distancing from the child. The frequency of burnout-related feelings is rated on a seven-point Likert scale ranging from “0” (never) to “6” (every day). Scores are calculated for both the overall scale and for each individual dimension. The tool demonstrates excellent internal consistency, with a Cronbach’s alpha of .96 (38).
5. **The perception of parenting experiences** will be measured using the Kansas Inventory of Parental Perceptions (KIPP, Behr et al., 1992; in the Polish adaptation: Pisula & Noińska, 2020). This tool consists of four scales: Positive child influence, Social comparisons, Search for cause, Sense of control. Participants respond to individual items within each scale by selecting one of four response options: “definitely not”, “no”, “definitely yes” and “yes”. Scores are calculated for each of the four scales. To date, no Polish studies have examined the factor structure, validity and reliability of the KIPP scales. In the original version, Cronbach’s alpha coefficients are .77 for the “Positive child contribution” scale, 0.66 for the “Social comparisons” scale, .87 for the “Search for cause” scale and .79 for the “Sense of control” scale (39).
6. **The family resilience** will be examined by the Family Resilience Assessment Scale (FRAS, Walsh, 1996; in the Polish adaptation: Nadrowska et al., 2021). This tool consists of 54 items across six subscales, which evaluate various aspects of family resilience. The subscales assess the following areas: Family communication and problem-solving, Use of social and economic resources, Maintaining a positive attitude, Family ties, Family spirituality, Ability to give meaning to adversity. Each item is rated on a four-point scale, with the following response options: “strongly agree”, “agree”, “disagree”, “strongly disagree”. Higher scores indicate higher level of family resilience. The internal reliability of the subscales is satisfactory, with a Cronbach’s alpha coefficient of.96 for the entire scale (40).

Furthermore, the following sociodemographic data will be collected:

1. mother’s age [in years],
2. education level [primary, secondary vocational, secondary technical, secondary general, higher bachelor’s degree, higher master’s degree],
3. place of living [village, town with a specified number of inhabitants],
4. marital status [single, married, informal union, divorced, separated, widow],
5. employment status [employed under an employment contract, employed under a civil law contract, own business, student, housewife, unemployed and looking for work, unemployed due to health condition, other],
6. economic situation [very good, good, average, poor, bad],
7. information on additional sources of income, benefits received [care benefits, emergency allowances, financial assistance from the family, foundation assistance, alimony, other],
8. estimated average monthly costs related to the child’s treatment and rehabilitation [in PLN],
9. number of children,
10. as well as details about the child with genetic disease [age, gender, birth order and specific genetic diagnosis].

These variables will help contextualize the findings and explore potential associations between sociodemographic factors and the levels of posttraumatic stress, posttraumatic growth, and other study outcomes.

### Sample size

To achieve the target sample size (N=120), participants will be recruited from various early intervention centers, hospitals, associations and foundations supporting parents and their children with genetic diseases. Recruitment will focus on centers providing treatment and rehabilitation for children in Poland. Given the assumption that 30% of participants may drop out by the second measurement, 40 participants will be recruited for each of the study groups:

- Group 1 – children aged from birth to 11 months 30 days,
- Group 2 – children aged from 12 months to 23 months 30 days,
- Group 3 – children aged from 24 months to 35 months 30 days,

(120 mothers in total). This will ensure 30 participants remain in each group by the second measurement, resulting in approx. 90 mothers completing the study.

## Data management

The data collected in the study, including information about both the mother and the child, will remain confidential. No personal data will be disclosed to the public. Only the researchers conducting the study will have access to the data, and the results will be presented in aggregate form.

## Statistical method

Sociodemographic data for the groups will be characterised by descriptive methods as: mean (SD), median (range), n (%). The results obtained in measurements I and II will be subjected to statistical analysis. The following statistical analyses are planned to be used: test of intergroup differences for dependent groups, test of intergroup differences for independent groups, Pearson r correlation, and regression analysis. If it is impossible to include the mother in measurement II, the mother will be excluded from the study. Results and conclusions will be developed based on statistical analyses.

## Discussion

We intend to explore whether the mother’s time perspective changes significantly during the experience of a child’s genetic disease. In this study, we take into account the periodization of child development. It included mothers of children with a genetic disease in early childhood, i.e., from birth to 3 years of age (41,42), as the sample homogenity allows for better generalization of the results. The sequential nature of the study allows for the observation of the dynamics of changes in posttraumatic stress and posttraumatic growth in mothers of children with genetic disease in a broader timeframe, while measurements from several time points are obtained at the same time.

The study will significantly contribute to the development of the discipline by undertaking the problem of the importance of time perception in the process of mothers’ coping with the situation of the child’s genetic disease. Although there are scientific reports on the role of time perspective on the intensity of posttraumatic stress and the level of posttraumatic growth, there is a gap in the scope of research on the relationship between the parent’s time perspective and the intensity of posttraumatic stress and the level of posttraumatic growth in the parent in the situation of the child’s genetic disease, while taking into account parental factors and the dynamics of changes occurring during the child’s disease. The posttraumatic stress rates studied in the group of mothers of children with genetic diseases will allow for quick intervention and providing the necessary psychological support to mothers by providing them with consultations and referring them to institutions providing specialist help.

The results of the study may constitute a solid theoretical basis for further experimental studies towards the use of elements of time perspective balancing therapy as a method of short-term therapeutic intervention for mothers in the first moments after receiving a diagnosis of a child’s genetic disease. Using mothers’ awareness of the impact of their time perspective on the ways of coping with a crisis situation may prove to be a potential therapeutic tool in clinical work.

However, the study has its limitations. Possible limitations of the study may include: a low retention rate and limited access to the study group. Analysis of the results will be possible if the mother undertakes to participate in both study measurements. If it is not possible to include the mother in the second measurement, the person will be excluded from the study. Due to the specificity of genetic diseases, which belong to the group of diseases with limited incidence, reaching the appropriate number of people studied may be difficult.

## Data Availability

No datasets were generated or analysed during the current study. All relevant data from this study will be made available upon study completion.

## Authors’ contributions

Conceptualization: Aleksandra Kołecka, łucja Bieleninik, Judyta Borchet Methodology: Aleksandra Kołecka, łucja Bieleninik, Judyta Borchet Supervision: łucja Bieleninik, Judyta Borchet

Writing– original draft: Aleksandra Kołecka

Writing– review & editing: Aleksandra Kołecka, łucja Bieleninik, Judyta Borchet

